# Glucocorticoids and B Cell Depleting Agents Substantially Impair Immunogenicity of mRNA Vaccines to SARS-CoV-2

**DOI:** 10.1101/2021.04.05.21254656

**Authors:** Parakkal Deepak, Wooseob Kim, Michael A. Paley, Monica Yang, Alexander B. Carvidi, Alia A. El-Qunni, Alem Haile, Katherine Huang, Baylee Kinnett, Mariel J. Liebeskind, Zhuoming Liu, Lily E. McMorrow, Diana Paez, Dana C. Perantie, Rebecca E. Schriefer, Shannon E. Sides, Mahima Thapa, Maté Gergely, Suha Abushamma, Michael Klebert, Lynne Mitchell, Darren Nix, Jonathan Graf, Kimberly E. Taylor, Salim Chahin, Matthew A. Ciorba, Patricia Katz, Mehrdad Matloubian, Jane A. O’Halloran, Rachel M. Presti, Gregory F. Wu, Sean P.J. Whelan, William J. Buchser, Lianne S. Gensler, Mary C. Nakamura, Ali H. Ellebedy, Alfred H.J. Kim

**Affiliations:** Inflammatory Bowel Diseases Center, Division of Gastroenterology, Department of Medicine, Washington University School of Medicine, St. Louis, MO, USA; Division of Immunobiology, Department of Pathology and Immunology, Washington University School of Medicine, St. Louis, MO, USA; Division of Rheumatology, Department of Medicine, Washington University School of Medicine, St. Louis, MO, USA; Division of Rheumatology, Department of Medicine, University of California San Francisco, San Francisco, CA, USA; Clinical Trials Unit, Washington University School of Medicine, St. Louis, MO, USA; Division of Gastroenterology, Department of Medicine, Washington University School of Medicine, St. Louis, MO, USA; Department of Genetics, Washington University School of Medicine, St. Louis, MO, USA; Department of Molecular Microbiology, Washington University School of Medicine, St. Louis, MO, USA; Department of Neurology, Washington University School of Medicine, St. Louis, MO, USA; Division of Infectious Diseases, Department of Medicine, Washington University School of Medicine, St. Louis, MO, USA; Inflammatory Bowels Diseases Center, Division of Gastroenterology, Department of Medicine, Washington University School of Medicine, St. Louis, MO, USA; The Andrew M. and Jane M. Bursky Center for Human Immunology and Immunotherapy Programs, Washington University School of Medicine, St. Louis, MO, USA; Arthritis/Immunology Section, San Francisco Veterans Administration Health Care System, San Francisco, CA, USA

## Abstract

**Background:** Individuals with chronic inflammatory diseases (CID) are frequently treated with immunosuppressive medications that can increase their risk of severe COVID-19. While novel mRNA-based SARS-CoV-2 vaccination platforms provide robust protection in immunocompetent individuals, the immunogenicity in CID patients on immunosuppression is not well established. Therefore, determining the effectiveness of SARS-CoV-2 vaccines in the setting of immunosuppression is essential to risk-stratify CID patients with impaired protection and provide clinical guidance regarding medication management.

**Methods:** We conducted a prospective assessment of mRNA-based vaccine immunogenicity in 133 adults with CIDs and 53 immunocompetent controls. Blood from participants over 18 years of age was collected before initial immunization and 1-2 weeks after the second immunization. Serum anti-SARS-CoV-2 spike (S) IgG^+^ binding, neutralizing antibody titers, and circulating S-specific plasmablasts were quantified to assess the magnitude and quality of the humoral response following vaccination.

**Results:** Compared to immunocompetent controls, a three-fold reduction in anti-S IgG titers (P=0.009) and SARS-CoV-2 neutralization (p<0.0001) were observed in CID patients. B cell depletion and glucocorticoids exerted the strongest effect with a 36- and 10-fold reduction in humoral responses, respectively (p<0.0001). Janus kinase inhibitors and antimetabolites, including methotrexate, also blunted antibody titers in multivariate regression analysis (P<0.0001, P=0.0023, respectively). Other targeted therapies, such as TNF inhibitors, IL-12/23 inhibitors, and integrin inhibitors, had only modest impacts on antibody formation and neutralization.

**Conclusions:** CID patients treated with immunosuppressive therapies exhibit impaired SARS-CoV-2 vaccine-induced immunity, with glucocorticoids and B cell depletion therapy more severely impeding optimal responses.

## INTRODUCTION

The Coronavirus disease 2019 (COVID-19) is a global pandemic caused by severe acute respiratory syndrome coronavirus-2 (SARS-CoV-2) that has infected millions, causing countless deaths and widespread economic devastation. Several vaccines against SARS-CoV-2 using either a novel liposomal mRNA-based delivery platform or an adenovirus-based approach have been authorized for emergency use by the Food and Drug Administration (FDA).^1-4^ The goal of vaccination is to generate long-lasting protection against infection. Most vaccines in clinical use achieve this protection at least in part through the generation of pathogen-specific antibody responses. Overcoming the COVID-19 pandemic will highly depend on the success of vaccine effectiveness.

The current management of various chronic inflammatory diseases (CID) including inflammatory bowel diseases (IBD), rheumatoid arthritis (RA), psoriasis, systemic lupus erythematosus (SLE), and multiple sclerosis (MS) typically requires immunosuppressive medications to achieve and maintain disease response and remission.^5-9^ Thus, patients with CID may be more vulnerable to infectious diseases. Indeed, certain medications such B cell depleting therapies (BCDT), glucocorticoids, and sulfasalazine have been associated with increased hospitalization and death due to COVID-19.^10,11^ Consequently, vaccination is recommended for CID patients. However, certain immunosuppressive medications have been shown to blunt vaccination responses.^12^

The seroconversion and magnitude of anti-SARS-CoV-2 antibody reactivity after COVID-19 infection have been reported to be attenuated in tumor necrosis factor-inhibitor (TNFi) [infliximab]-treated IBD patients compared with a gut-selective anti-integrin (vedolizumab), which is further blunted with concomitant thiopurine or methotrexate.^13^ This suggests that other immunosuppressives may also attenuate humoral responses following SARS-CoV-2 vaccination. The novel delivery platform (liposomal mRNA), along with the lack of data on efficacy and safety in immunosuppressed subjects due to exclusion from clinical trials,^1,2^ has led to uncertainty whether to continue or hold immunosuppression to maximize SARS-CoV-2 vaccine efficacy, and discordant guidelines from national medical organizations.^14-16^

Emerging data demonstrate reduced antibody responses in immunosuppressed individuals following mRNA vaccination. Organ transplant recipients receiving antimetabolite therapy and older recipients were less likely to develop an antibody response following the first dose of the BNT162b2 or the mRNA-1273 vaccines.^17^ Furthermore, CID patients (n=26) vaccinated with mRNA vaccines had blunted anti-S IgG levels with modest reduction in neutralization in ∼25% of subjects compared to immunocompetent controls.^18^ Similarly, in 13 IBD patients treated with either TNFi or anti-integrin therapy who completed the two-dose vaccine schedules, reductions in anti-S IgG levels were observed compared to non-IBD controls.^19^ While these early observations corroborate suspicions that immunosuppression can reduce antibody responses following SARS-CoV-2 vaccination, these studies have inadequate sample sizes to determine which specific immunosuppressive classes drive a reduction in humoral responses to SARS-CoV-2 vaccination.

Here, we report immunogenicity data in 133 patients with CID and 53 immunocompetent controls following completion of the two-dose mRNA SARS-CoV-2 vaccination series. The size of our cohort enables us to test the hypothesis that certain immunosuppressive therapies mediate reductions in vaccine-induced humoral immune responses.

## METHODS

### Study design

The COVaRiPAD (**CO**VID-19 **Va**ccine **R**esponses **i**n **P**atients with **A**utoimmune **D**isease) study is a longitudinal observational study seeking to elucidate the magnitude, quality, and evolution of the immune response to SARS-CoV-2 vaccines. The initial phase of COVaRiPAD specifically examined the magnitude and quality of the acute humoral response.

### Patient recruitment

133 participants with confirmed CID and 53 immunocompetent controls recruited from the faculty, employees, staff, and patients at Washington University School of Medicine and BJC Healthcare system (St. Louis, MO, USA) and the University of California, San Francisco, UCSF Health, and Zuckerberg San Francisco General Hospital (San Francisco, CA, USA) from 12/10/2020 to 3/20/2021. All participants provided written informed consent. Participants were assessed within two weeks prior to initial vaccination and twenty days post-final vaccination, with 96% of blood samples collected within 14 days post-vaccination. Inclusion and exclusion criteria for CID and immunocompetent controls can be found in the Supplemental Appendix. Medications, including dose and last administration date were confirmed at each study visit. Medications comprising antimetabolites and BCDT are listed in the Supplemental Appendix. All patients continued their immunosuppressive medications per their treating physician, except for five individuals who held methotrexate around immunization.

### Assessment of humoral responses

As previously described,^20^ anti-S IgG quantification was performed using enzyme-linked immunosorbent assay (ELISA), and direct *ex-vivo* enzyme-linked immunosorbent spot (ELISpot) assays were performed to quantify recombinant S protein-binding IgG-secreting cells. Neutralization assays were performed using a fluorescence-based platform leveraging a chimeric vesicular stomatitis virus (VSV) pseudotyped with the common variant strain (D614G) of the S protein of SARS-CoV-2 that has been modified for high-throughput processing.^21,22^ Additional details are provided in the Supplementary Appendix.

### Statistical analysis

Statistical analyses were performed using Prism v9.1.0 (GraphPad Software) and Stata/MP 13.1. Kruskal-Wallis test followed by Dunn’s multiple comparison’s test or Mann-Whitney test were used as indicated. Effects were further refined via tobit linear regression to account for left-censoring of data below the limit of detection (LoD) and to adjust for pre-vaccination titer, age, and gender. For regression analysis, to test for the effects of medications in the presence of combination therapy, medications were ranked by their impact on vaccine response and patients included in discrete groups by medication having the largest effect size.

### Ethics approval

This study was approved by the Washington University School of Medicine Institutional Review Board (protocol #201105110, approved 6/1/2011; protocol #202012081, approved 12/21/2020; and protocol #202012084, approved 12/23/2020) and the UCSF Institutional Review Board (protocol #17-21898, approved 4/22/2017 and protocol #20-33078, approved 1/4/2021).

## RESULTS

Between December 2020 and March 2021 at two sites (St. Louis, MO and San Francisco, CA), 133 patients with CIDs and 53 controls were enrolled. The mean age was 45.5±16.0 years old with 14.3% >65 years old (**Table 1, Table S1**). The majority were female (74.4%) and white (88.0%). The most common CID diagnoses among the subjects were 31.6% with IBD and 28.6% RA. The most common immunosuppressive medications were TNFi (28.6%) and methotrexate (21.8%).

**Table 1.** Demographic and Clinical Characteristics of Participants with Chronic Inflammatory Diseases.

| Characteristics | N=133 |
| --- | --- |
| Age, years<br>Mean ± SD | 45.5 ± 16.0 |
| Age category — no. of participants (%) |  |
| <65yr | 114 (85.7) |
| ≥65yr | 19 (14.3) |
| Gender — no. of participants (%)<sup>‡</sup> |  |
| Female | 99 (74.4) |
| Male | 34 (25.6) |
| Hispanic or Latinx ethnicity – no of participants (%)<sup>‡</sup> |  |
| Hispanic or Latinx | 6 (4.5) |
| Not Hispanic or Latinx | 127 (95.5) |
| Race or ethnic group — no. of participants (%)<sup>‡</sup> |  |
| White | 117 (88.0) |
| Asian | 9 (6.8) |
| Black or African American | 4 (3.0) |
| American Indian or Alaska Native | 2 (1.5) |
| Multi-racial | 1 (0.8) |
| BMI [kg/m<sup>2</sup>], mean ± SD | 26.6 ± 6.3 |
| Days after 2nd immunization for blood sample, mean ± SD | 8.5 ± 2.8 |

| Immunologic Diagnosis - no. of participants (%)<sup>§</sup> |  |
| --- | --- |
| Inflammatory Bowel Disease | 42 (31.6) |
| Crohn's Disease | 22 (16.5) |
| Ulcerative Colitis | 18 (13.5) |
| Other | 2 (1.5) |
| Rheumatoid Arthritis | 38 (28.6) |
| Spondyloarthritis | 20 (15) |
| Axial Spondyloarthritis | 6 (4.5) |
| Psoriatic Arthritis/Psoriasis | 10 (7.5) |
| IBD Arthritis | 4 (3.0) |
| Uveitis | 5 (3.8) |
| Systemic Lupus Erythematosus | 15 (11.3) |
| Other Connective Tissue Disease<sup>¶</sup> | 4 (3.0) |
| Sjögren's Syndrome | 8 (6.0) |
| Vasculitis | 5 (3.8) |
| Autoinflammatory Syndrome | 2 (1.5) |
| Multiple Sclerosis | 9 (6.8) |
| Neuromyelitis Optica | 1 (0.8) |
| IgG4-Related Disease | 2 (1.5) |
| Hidradenitis Suppurativa | 1 (0.8) |
| Human Immunodeficiency Virus | 1 (0.8) |
| Anti-phospholipid Syndrome | 1 (0.8) |
| **Medication exposure - no. of participants (%)** |  |
| Prednisone | 17 (12.8) |
| Mean mg/day ± SD | 6.5 ± 5.8 |
| Range, mg/day | 1 - 20 |
| Disease Modifying Antirheumatic Drug (DMARD) |  |
| Methotrexate | 29 (21.8) |
| Mean mg/week ± SD | 17.1 ± 5.4 |
| Range, mg/day | 7.5 - 25 |
| Hydroxychloroquine | 30 (22.6) |
| Mycophenolate Mofetil | 9 (6.8) |
| Azathioprine | 4 (3.0) |
| Leflunomide | 2 (1.5) |
| Sulfasalazine | 7 (5.3) |
| Janus Kinase inhibitors |  |
| Tofacitinib | 10 (7.5) |
| Upadacitinib | 1 (0.8) |
| Biological therapies |  |
| Tumor Necrosis Factor inhibitors<sup>§</sup> | 38 (28.6) |
| B cell depleting therapies<sup>†</sup> | 10 (7.5) |
| Belimumab | 3 (2.3) |
| Vedolizumab | 12 (9.0) |
| Interleukin 12/23 or 23 inhibitors* | 10 (7.5) |
| Abatacept | 2 (1.5) |
| Tocilizumab | 1 (0.8) |
| Canakinumab | 1 (0.8) |
| Fingolimod | 1 (0.8) |
| Ibrutinib | 1 (0.8) |
| Nonsteroidal Anti-inflammatory Drugs (NSAIDs) | 27 (20.3) |
| No DMARDs or biologics | 9 (6.8) |

<sup>§</sup>Patients could be diagnosed with more than 1 condition, such that the sum of diagnoses is greater than the number of participants.
<sup>‡</sup>Gender, Race or ethnic group was reported by the participant.
<sup>¶</sup>Other Connective Tissue Disease includes Undifferentiated connective tissue disease and Mixed Connective Tissue Disease.
<sup>§</sup>Tumor Necrosis Factor Inhibitors includes adalimumab (n=13), certolizumab pegol (n=5), etanercept (n=13), golimumab (n=2) and infliximab (n=6).
<sup>†</sup>B cell depleting therapies include rituximab (n=8) and ocrelizumab (n=4).
\*Interleukin 12/23 inhibitors include ustekinumab (n=9) and interleukin 23 inhibitors include guselkumab (n=1)

Immunocompetent controls and most CID participants developed robust anti-SARS-CoV-2 antibody responses after immunization, although CID participants averaged a 3-fold reduction in antibody titers compared to immunocompetent controls (**Figure 1A-B**). This diminished antibody response was associated with a similar 2.7-fold decrease in neutralization of viral infection **(Figure 1C)**. Additionally, we observed a non-statistically significant trend towards impaired plasmablast formation in CID participants (**Figure S1A**).

**Figure 1.**
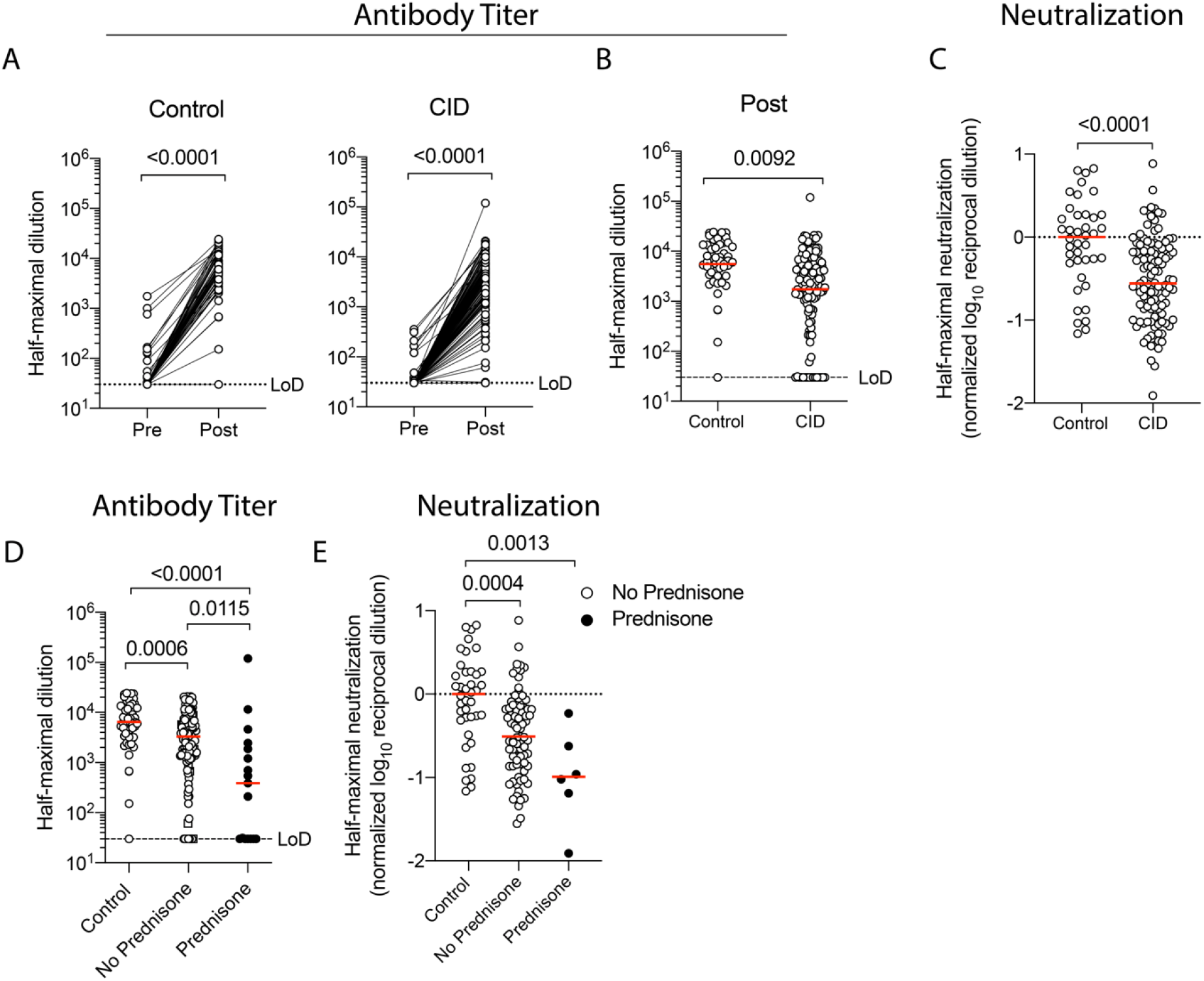
Glucocorticoids Reduce Immunogenicity of mRNA-based SARS-CoV-2 Vaccination. Immunocompetent (control) and chronic inflammatory disease (CID) participants underwent blood draws pre- and post-vaccination (∼1-2 weeks after boost) and concurrent medication use was collected. Quantification of circulating anti-S IgG for immunocompetent (left) and CID (right) participants pre- and post-immunization is shown in Panel A. Each symbol represents an individual for each timepoint. Dotted lines indicate limits of detection (LoD). Horizontal red lines indicate median value. A lower titer of anti-SARS-CoV-2 antibodies post-vaccination in CID vs immunocompetent participants is shown in Panel B. Neutralization of SARS-CoV-2 S protein by serum of immunocompetent and CID participants post-vaccination is shown in Panel C. Pseudotyped vesicular stomatitis virus with SARS-CoV-2 S protein was introduced to Vero cells with decreasing concentrations of serum to identify the dilution required for 50% neutralization. Values are normalized to the median of immunocompetent participants. A lower titer of anti-SARS-CoV-2 antibodies is found in CID participants taking glucocorticoids is shown in Panel D. CID participants are split by prednisone use (shaded black). A decrease in neutralization of SARS-CoV-2 S for CID participants on or off glucocorticoids is shown in Panel E. P values are indicated for each comparison (Dunn’s multiple comparison test).

To identify which medications may reduce the immunogenicity of mRNA-based SARS-CoV-2 vaccination, we first compared participants on or off glucocorticoids with immunocompetent controls. Glucocorticoid use resulted in a 10-fold reduction in anti-S IgG and neutralization titers, as well as fewer circulating plasmablasts after vaccination (**Figure 1D-E, Figure S1B)**. Strikingly, seropositivity post-vaccination decreased from 98% in immunocompetent controls, to 92% in CID participants off prednisone, to 65% in CID participants on prednisone. Thus, the use of glucocorticoids negatively impacts the immunogenicity of mRNA-based vaccines.

We next examined whether antimetabolites, particularly methotrexate, influenced the immunogenicity of mRNA-based vaccination in the absence of glucocorticoids. Compared to immunocompetent controls, CID participants on any antimetabolite demonstrated on average a 2-3-fold reduction in anti-spike antibodies and neutralization **(Figure 2A-B)**. This effect was seen when specifically examining the effect of methotrexate. We also observed a small decline in circulating plasmablasts associated with antimetabolites, but this sub-analysis contained a high degree of variance **(Figure S2)**. Thus, antimetabolite therapy, including methotrexate, modestly impairs the immunogenicity of mRNA-based vaccines.

**Figure 2.**
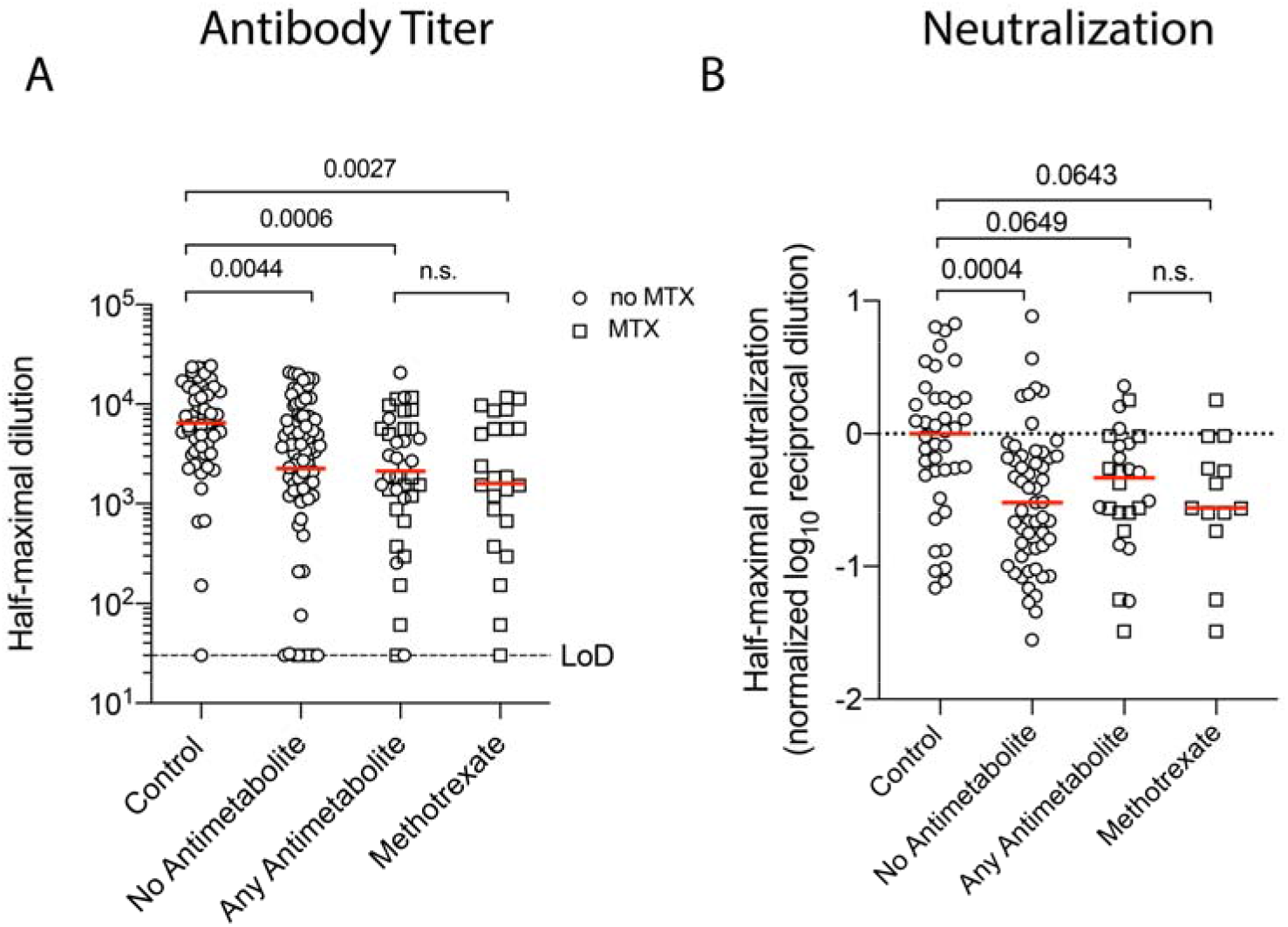
Antimetabolites Modestly Reduce Immunogenicity of mRNA-based SARS-CoV-2 Vaccination. Decreases in circulating anti-S IgG post-immunization for CID participants either not taking antimetabolites, taking any antimetabolite, or specifically taking methotrexate are shown in Panel A. Methotrexate use is denoted by square symbols. Glucocorticoid use was excluded. Dotted lines indicate limits of detection (LoD). A decrease in neutralization of SARS-CoV-2 S for CID participants on or off antimetabolites is shown in Panel B. Values are normalized to the median of immunocompetent participants. Each symbol represents an individual for each timepoint. Horizontal red lines indicate median value. P values are indicated for each comparison (Dunn’s multiple comparison test).

Given previous reports of targeted therapies impairing vaccine responses,^12^ we examined the impact of multiple agents on mRNA-based vaccination. Compared to immunocompetent controls, we noted a reduction in antibody titers, neutralization, and circulating plasmablasts for CID participants on TNFi agents; however only neutralization reached statistical significance **(Figure 3A-B, Figure S3)**. We did not observe lower anti-S IgG titers in participants taking a combination of TNFi with methotrexate compared to TNFi alone **(Figure 3C-D)**.

**Figure 3.**
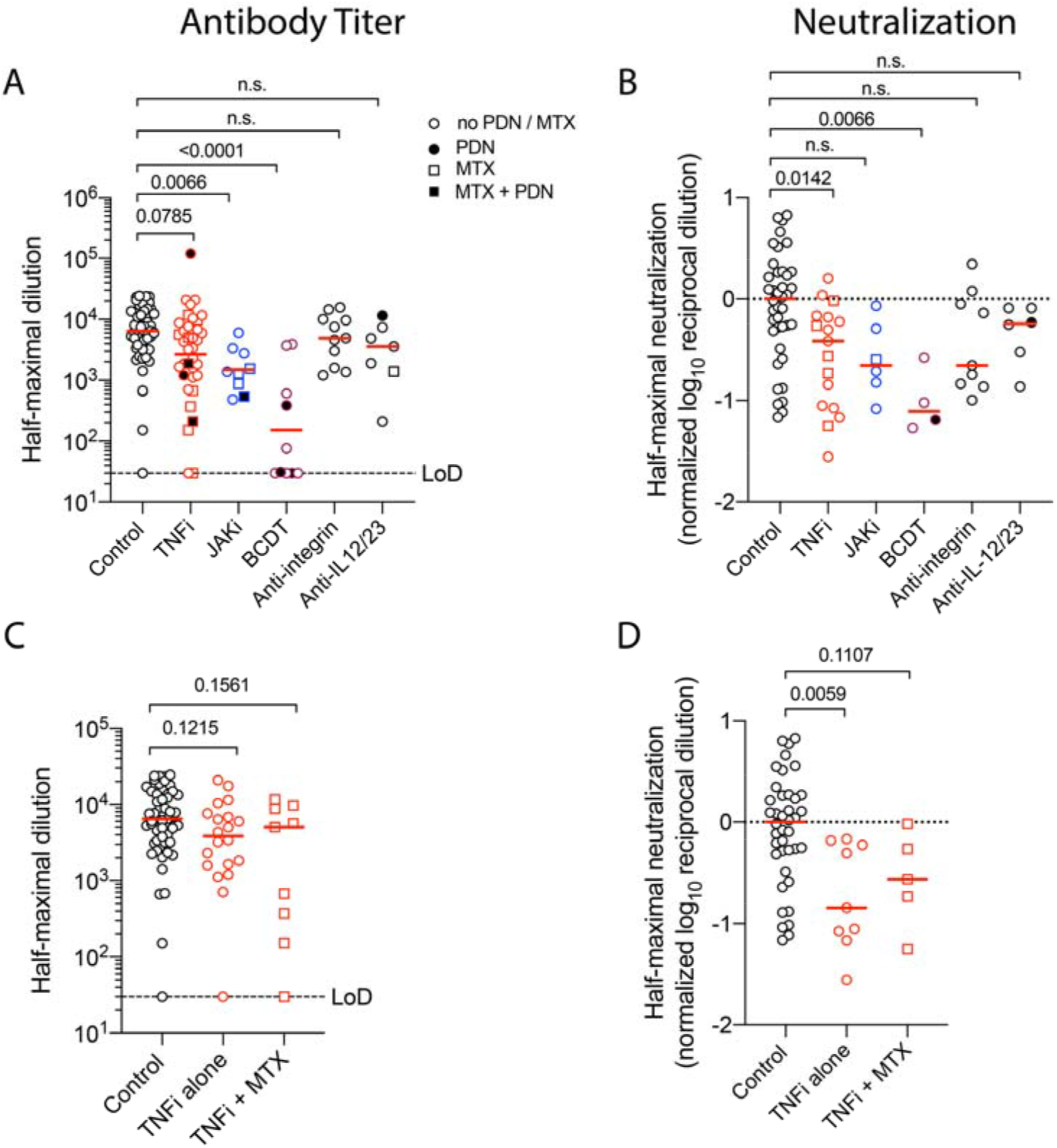
B Cell Depletion Therapy Reduces Immunogenicity of mRNA-based SARS-CoV-2 Vaccination. Quantification of circulating anti-S IgG for immunocompetent (control) and CID participants on TNF inhibitors (TNFi), JAK inhibitors (JAKi), B cell depletion therapy (BCDT), anti-integrin agents, and anti-IL-12/23 agents are shown in Panel A. Combined use with prednisone (PDN) is denoted by solid fill and with methotrexate (MTX) is denoted by square symbols. Dotted lines indicate limits of detection (LoD). Neutralization for the same sub-groups is shown in Panel B. Values are normalized to the median of immunocompetent participants. Quantification of circulating anti-S IgG for CID participants on TNFi, separated by methotrexate use is shown in Panel C. A reduction in neutralization for participants on an TNFi alone is shown in Panel D. Each symbol represents an individual for each timepoint. Horizontal red lines indicate median value. P values are indicated for each comparison (Dunn’s multiple comparison test).

JAKi use also drove a reduction in anti-S IgG and neutralization, although the latter did not reach statistical significance **(Figure 3A-B)**. In contrast to TNFi and JAKi, BCDT were associated with a meaningful 36-fold reduction in anti-S IgG and neutralization titers, as well as absence of circulating plasmablasts in one subject where data was available **(Figure 3A-B, Figure S3)**. The reduction in seroconversion was most striking in participants who had more recently (within 6 months) received BCDT (**Figure S4A**) with variable recovery over time (**Figure S4B)**. Other targeted therapies were associated with only a modest reduction in antibody titers, neutralization, or circulating plasmablasts that did not reach statistical significance **(Figure 3A-C, Figure S3)**.

To test the strength of association between medications and reduction in antibody titers, regression analyses were performed. We found that, when compared to immunocompetent participants, CID participants on BCDT, prednisone, JAKi, and antimetabolites all had statistically significant reductions in antibody titers in univariate and multivariate models (**Table 2, Figure S5**). In contrast, antimalarials (i.e., hydroxychloroquine) and TNFi were not significantly associated with reduced antibody titers. This effect of BCDT, prednisone, and JAKi, but not antimetabolites, remained significant when compared to CID participants who were not taking BCDT, prednisone, JAKi, antimetabolites, antimalarials, or TNFi **(Table S2**). Furthermore, there was no interaction between TNFi and methotrexate therapy in the reduction of antibody titers beyond the effect of methotrexate alone. In contrast, participants taking multiple medications displayed an 83.1% reduction in antibody titers compared to those not on prednisone **(Table S3)**.

**Table 2:** Post-vaccine Titer by Chronic Inflammatory Diseases Medication Group versus Immunocompetent Participants.

| **Medication group<sup>1</sup>** | **N** | **Fold Reduction in Titer (95% CI)** | **P value<sup>2</sup>** |
| --- | --- | --- | --- |
| B-cell depleting therapy | 10 | 52.7 (18.3 - 152.0) | **4.0E-10** |
| Prednisone | 14 | 15.1 (4.9 - 46.3) | **9.0E-06** |
| JAK inhibitors | 10 | 6.6 (2.9 - 15.3) | **3.0E-05** |
| Antimetabolites | 31 | 2.9 (1.5 - 5.6) | **0.0023** |
| Antimalarials | 14 | 2.2 (0.96 - 5.0) | 0.062 |
| TNF inhibitors | 21 | 1.7 (0.92 - 3.2) | 0.091 |
| Other<sup>3</sup> or No Medications | 33 | 1.6 (0.92 - 2.8) | 0.097 |

<sup>1</sup>Each medication group excludes patients in the group(s) above.
<sup>2</sup>Tobit regression of $\log_{10}$ (post-vaccine titer) left-censored at limit of detection $\log_{10}(30)$ , adjusting for $\log_{10}$ (pre-vaccine titer), age, and gender.
<sup>3</sup>Other medications include NSAIDs, sulfasalazine, anti-integrin therapy, anti-IL-12/23 therapy, and ibrutinib.

## DISCUSSION

We report the analysis from the initial phase of the COVaRiPAD study, focusing on antibody responses after two doses of either BNT162b2 or mRNA-1273. Using a cohort composed of 133 patients with CID and 53 immunocompetent controls recruited from two tertiary medical centers in the United States, we show that CID patients have a greater reduction in magnitude and quality of antibody responses compared to immunocompetent controls. This is consistent with published or preprint reports in patients with CID.^18,19,23^ Furthermore, BCDT, glucocorticoids, JAKi, and, to a lesser extent, antimetabolites were associated with reduced antibody responses.

Patients on BCDT had substantial reductions in anti-S IgG as well as neutralizing antibodies, as was observed for influenza and pneumococcal vaccination.^24,25^ We found this was most prominent when vaccination occurred within 6 months of BCDT administration, with gradual recovery after 9 months post-treatment with rituximab. All four subjects on ocrelizumab had undetectable antibodies, and it remains unclear whether they will experience similar recovery kinetics as those on rituximab.

Ocrelizumab has improved antibody-dependent cellular cytotoxicity compared to rituximab, which may prolong its impact on SARS-CoV-2 immunization.^26^ Consistent with our data, after immunization with BNT162b2, another patient with MS failed to generate detectable anti-S antibodies six months after ocrelizumab treatment.^27^ These data are concerning as COVID-19 patients on BCDT have increased mortality compared to methotrexate users.^11^ Notably, in the VELOCE study, immunization against keyhole limpet hemocyanin (KLH), influenza, pneumococcus, and tetanus toxoid elicited antibody responses, albeit reduced, despite ocrelizumab administration 12 weeks prior. ^28^ Importantly, the KLH and influenza vaccinations consist of three immunization series, raising the question of whether additional boosters in this population may be needed to mount responses. Thus, it may be beneficial to withhold BCDT for some duration or, after appropriate study, provide an additional vaccine booster to enable reasonable antibody responses following mRNA-based SARS-CoV-2 vaccination.

We also found that glucocorticoid use resulted in a 10-fold reduction in antibody titers following SARS-CoV-2 vaccination. This was independent of dose, with very low titers observed in some subjects taking <5mg prednisone daily. Furthermore, use of prednisone in combination with other medications further attenuated antibody titers compared to other medications taken alone. In SLE subjects, high dose prednisone use (>20mg daily) was associated with a decline in seroconversion after influenza vaccination, but a dose-independent effect on seroconversion was observed following pneumococcus, tetanus toxoid, *Haemophilus influenzae* type B, or hepatitis B vaccination.^29-31^ In contrast, glucocorticoids had minimal impact on seroconversion after influenza vaccination, either at high doses in the setting of asthma or low doses in RA or spondylarthritis patients.^32,33^ Importantly, repeated immunization against hepatitis B can still boost suboptimal humoral response in immunosuppressed individuals.^34^ Thus, mRNA-based SARS-CoV-2 vaccination may be more sensitive to the immunosuppressive effects of glucocorticoid use than traditional platforms, and additional booster immunizations may be required in this population to achieve protection. As most prednisone users were on other immunosuppressants in this study, efforts to taper prednisone as safely as possible while initiating additional therapies may be needed to permit optimal antibody responses from mRNA-based vaccines.

The use of methotrexate, TNFi, and JAKi all lowered both antibody and neutralization titers, but only JAKi and antimetabolites demonstrated a statistically significant association in a regression model. Similar data for reduced antibody titers in TNFi users was also noted in the IBD population.^23^ Reassuringly, the combination of methotrexate with TNFi did not seem to further reduce antibody titers relative to each agent alone. Furthermore, patients who have not held antimetabolite, TNFi, or JAKi therapy can be reassured that the impact on antibody titers is modest.

IL-12/23 inhibitors and vedolizumab had minimal impact on antibody titers. In contrast, lower anti-S titers have been reported after SARS-CoV-2 vaccination with vedolizumab.^19^ We noted a reduction of neutralization titers for those on vedolizumab, but the low sample size may have limited the ability to detect a significant effect. Additional recruitment will be needed to confirm this observation.

Prior to any evaluation of mRNA-based vaccines in CID patients, professional societies were pressed to provide guidance regarding the use of immunosuppression in CID patients. Current guidelines in IBD and psoriasis do not suggest holding biologics, small molecules, or antimetabolites prior to vaccination against SARS-CoV-2.^14,16^ In contrast, the COVID-19 Vaccine Clinical Guidance Summary from the American College of Rheumatology (ACR) has a moderate-level consensus in holding certain immunosuppressants such as methotrexate and JAKi for 1 week after each vaccine dose for those with well-controlled disease.^15^ The blunting of humoral responses in the setting of antimetabolites, TNFi, and JAKi support the initial recommendations from the ACR that certain medications may need to be held in order to optimize vaccine response. Importantly, our study was conducted prior to the release of these recommendations and only five subjects in our cohort held any medications (all methotrexate).

As no cutoff titer has been defined to best associate with protection, the impact of reduced antibody levels on protection remains unclear. Additionally, these data do not directly evaluate protection from SARS-CoV-2 infection nor prevention of hospitalization. Nevertheless, S-specific antibody titers in CID patients (particularly TNFi and JAKi) are comparable to patients with rapid recovery from COVID-19 and may, therefore, provide sufficient humoral protection.^35^ Furthermore, CD4^+^ and CD8^+^ T cell responses were observed in patients with COVID-19,^36^ suggesting the potential importance of T-cell-mediated immunity against SARS-CoV-2. Indeed, S-specific T cells are easily detected in those previously infected with SARS-CoV-2 immunized with a single dose of either mRNA-based vaccine.^37^ We plan to examine T cell responses in our cohort to determine this important aspect of post-vaccinal immunity to SARS-CoV-2.

The limitations of the study include small sample sizes for several classes of immunosuppressives (i.e. anti-IL-17, belimumab, abatacept), preventing any analysis. Additionally, we cannot evaluate differences between the two mRNA vaccines, nor with the adenovirus delivery platform. We also have limited racial and ethnic diversity due to selection bias (most subjects are faculty/staff at academic teaching hospitals). We have continued recruitment to address this. We will report safety, reactogenicity, disease state, and changes in disease activity in future studies.

Assessing long-term responses (i.e. 6 months post-booster) and neutralization of variants of concern will be critical as the impact of immunosuppression on vaccine response durability and epitope breadth is unknown. Most immunocompetent subjects vaccinated with mRNA-1273 only had modest reductions in neutralization 3 months post-booster,^38^ while recovered COVID-19 patients, maintained antibody, B, and T cell responses up to 8 months after infection.^39^ Of concern though, is the poor neutralization responses to B.1.351 in previously uninfected immunocompetent subjects following mRNA vaccination.^40^ Whether immunosuppression may further attenuate cross-variant neutralization is unclear.

In summary, this initial analysis of the COVaRiPAD study, focusing on the magnitude and quality of antibody responses after two doses of either BNT162b2 or mRNA-1273, reveals that most patients with CID on immunosuppressive treatment were able to mount responses. Select classes, including JAKi, TNFi, methotrexate, glucocorticoids, and particularly BCDT have a varying deleterious impact on the magnitude and quality of anti-spike IgG and neutralizing responses compared to immunocompetent controls. These data serve as the initial building block for providers and professional societies to reach a broader consensus regarding the use of specific immunosuppressive agents in the peri-vaccination period.

## Data Availability

Relevant data are available from the corresponding author upon reasonable request.

## ACKNOWLEDGEMENTS

The authors would like to thank the study participants for their generous and enthusiastic participation, the treating clinicians for their support of the study recruitment process, and the staff of the Washington University School of Medicine Infectious Diseases Clinical Research Unit, Clinical Trials Research Unit, Center of Clinical Studies, and the Digestive Diseases Research Core Center for assistance with study visits. We thank the Siteman Tissue Procurement Core for processing and storage of the blood specimens. We thank Grace L. Paley for constructive comments on the manuscript. A.H.J.K. thanks Jinoos Yazdany for constructive conversations.

## Contributors

A.H.J.K., R.M.P., P.D., M.A.C, G.W., and A.H.E. conceived and designed the study. P.D., W.K., M.A.P., and A.H.J.K. composed the manuscript. A.E-Q., L.E.M., R.S., M.K., B.D.N., P.K., J.A.O., R.M.P., and A.H.J.K wrote and maintained the IRB protocols. P.D., M.A.P., A.C., A.E-Q., M.G., S.A., J.G., A.H., K.H., B.K., L.E.M., D.P., D.C.P., M.A.C., G.F.W., S.C., and R.S. recruited subjects, and A.C., A.E-Q., A.H., K.H., B.K., L.E.M., D.P., S.E.S., and D.C.P. phlebotomized participants and coordinated sample collection. W.K., M.T., M.J.L., and Z.L. performed experiments. P.D., W.K., M.A.P., K.E.T., W.J.B., L.S.G., M.C.N., A.H.E., and A.H.J.K. analyzed the data. M.C.N., M.M., and L.S.G. led formation, sample and data collection of UCSF cohort. All authors reviewed the manuscript.

