## Supplemental for "Glucocorticoids and B Cell Depleting Agents Substantially Impair Immunogenicity of mRNA Vaccines to SARS-CoV-2"

**SUPPLEMENTARY APPENDIX**

### COVaRiPAD Study Full Roster

Suha Abushamma, M.D., Seweuse Akuse, M.P.H., Megan Arb, B.S., Teresa Arb, B.S.N., R.N., C.C.R.C., Joel Brune, R.N., B.S.N., Jennifer Bruns, R.N., William J. Buchser, Ph.D., Samantha L. Burdess, M.S. Alexander B. Carvidi, B.A., Salim Chahin, M.S.C.E., M.D., Matthew A. Ciorba, M.D., PeChaz L. Clark, B.A., Rachel Cody, B.S.N., R.N., C.C.R.C., Parakkal Deepak, M.B.B.S, M.S., Ali H. Ellebedy, Ph.D., Alia A. El-Qunni, B.A., Lacey Feigl, H.S., Lianne S. Gensler, M.D., Maté Gergely, M.D., Jonathan Graf, M.D., Alem Haile, B.S., Katherine (Lulu) Huang, B.A., Patricia Katz, Ph.D., Alfred H.J. Kim, M.D., Ph.D., Wooseob Kim, Ph.D., Baylee Kinnett, B.S., Michael Klebert, Ph.D., R.N., A.N.P-B.C., Mariel J. Liebeskind, B.A., Zhuoming Liu, Ph.D., Mehrdad Matloubian, M.D., Lily E. McMorrow, B.A., Lynne Mitchell, M.A., M.S., Mary Nakamura, M.D., Tina Nolte, B.A., Darren Nix, H.S., Jane A. O’Halloran, M.D, Ph.D., Diane Paez, B.S., Michael A. Paley, M.D., Ph.D., Dana C. Perantie, M.P.H., Mackenzie Poole, B.A., Rachel M. Presti, M.D, Ph.D., Liz Reimer, R.N., B.S.N., Abbey Rose, H.S., Shea M. Roesel-Wakeland, B.A., Rebecca E. Schriefer, B.A., Shannon E. Sides, B.A., Kelly Streckfuss, R.N., B.S.N., C.C.R.C., Alyssa M. Spurling, M.A., Kimberly E. Taylor, Ph.D., M.P.H., Mahima Thapa, B.S., Sean P.J. Whelan, Ph.D., Gregory F. Wu, M.D., Ph.D., Monica Yang, M.D., Brittany Zwijack, R.N., B.S.N., C.C.R.C.

### Inclusion and Exclusion Criteria

**Subject Inclusion Criteria**

1. Able to understand and give informed consent.
2. Capable of attending all mandatory study visits according to the study schedule.
3. Males or females between over age 18.
4. For the chronic inflammatory disease (CID) cohort, have health care provider-documented CID. For the immunocompetent controls, must in good health as determined by medical history and physical exam and not be on any immunomodulatory nor immunosuppressive medications.

**Subject Exclusion Criteria**

1. History or prior SARS-CoV-2 vaccination or participation in an investigational study of SARS-CoV2 vaccines in the past two years. Of note, history of prior suspected or confirmed SARS-CoV-2 infection is not exclusionary.
2. Any history of allergy to vaccination.
3. Any history of Guillain-Barre post vaccination.
4. Have an acute illness or fever within 72 hours before vaccination.
5. A history of uncontrolled HIV infection or cancer (particularly leukemia, lymphoma, use of antineoplastic drugs or X-ray treatment). Persons with previous skin cancers or cured non-lymphatic tumors are not excluded from the study. HIV infection is considered controlled if there is documentation of stable antiretroviral regimen for the past 6 months and current CD4 count is > 300 with undetectable viral load.
6. History of any chronic medical conditions that are considered progressive or uncontrolled and have required hospitalization in the past 3 months (e.g., diabetes, heart disease, lung disease, liver disease, kidney disease, and uncontrolled hypertension).
7. History of excessive alcohol consumption, drug abuse, psychiatric conditions, social conditions or occupational conditions that in the opinion of the investigator would preclude compliance with the study.
8. Recipient of a blood products within 90 days of the vaccination visit, excluding intravenous immunoglobulin.
9. Have received any licensed live vaccine within 30 days or any licensed inactivated vaccine within 14 days prior to SARS-CoV-2 vaccination.
10. Have planned vaccination with any other vaccine during first 60 days of study participation.
11. Have donated blood or blood products within 30 days before study vaccination, plan to donate blood at any time during the duration of subject study participation, or plan to donate blood within 30 days after the last blood draw.
12. Any condition in the opinion of the investigator that would interfere with the proper conduct of the trial.

### Detailed Methods

**Classification of medications** Methotrexate, leflunomide, azathioprine, and mycophenolate mofetil were classified as antimetabolites, while rituximab and ocrelizumab were categorized together as BCDT.

**Sample collection and storage**

Serum, plasma, and peripheral blood mononuclear cells (PBMCs) were collected using Vacutainer CPT tubes (BD). Serum and plasma were immediately used or frozen at -80°C. PBMCs isolated using Ficoll density gradient centrifugation were immediately used or cryopreserved in 10% dimethylsulfoxide in fetal bovine serum (FBS).

**ELISA** Assays were performed in 96-well plates (MaxiSorp; Thermo) coated with 100 μl of recombinant spike (S) protein diluted to 1 μg/ml in PBS, and plates were incubated at 4°C overnight. Plates were then blocked with 10% FBS and 0.05% Tween20 in PBS. Serum or plasma were serially diluted in blocking buffer and added to the plates. Plates were incubated for 90 min at room temperature and then washed 3 times with 0.05% Tween-20 in PBS. Goat anti-human IgG-HRP (Jackson ImmunoResearch, 1:2,500) was diluted in blocking buffer before adding to wells and incubating for 60 min at room temperature. Plates were washed 3 times with 0.05% Tween-20 in PBS, and then washed 3 times with PBS before the addition of peroxidase substrate (SigmaFAST o-Phenylenediamine dihydrochloride, Sigma-Aldrich). Reactions were stopped by the addition of 1 M hydrochloric acid. Optical density measurements were taken at 490 nm. The half-maximal binding dilution for each serum or plasma sample was calculated using nonlinear regression (GraphPad Prism v9). The limit of detection was defined as 1:30.

**ELISpot** Direct ex-vivo ELISpot was performed to determine the number of total vaccine-binding, or recombinant S-binding IgG-secreting cells present in PBMC samples using IgG/IgA double-color ELISpot Kits (Cellular Technology Limited) according to the manufacturer’s instructions. Plates were coated overnight at 4°C with Flucelvax Quadrivalent 2019/2020 seasonal influenza virus vaccine (diluted 1:100), and 5 μg/ml recombinant S proteins, anti-human Ig. ELISpot plates were analyzed using an ELISpot counter (Cellular Technology Limited)

**Antigens** Recombinant soluble SARS-CoV-2 S protein were expressed as previously described.^1^ Briefly, mammalian cell codon-optimized nucleotide sequence coding for the soluble ectodomain of the S protein of SARS-CoV-2 (GenBank: MN908947.3, amino acids 1-1213) including a C-terminal thrombin cleavage site, T4 foldon trimerization domain, and hexahistidine tag was cloned into mammalian expression vector pCAGGS. The S protein sequence was modified to remove the poly basic cleavage site (RRAR to A) and two stabilizing mutations were introduced (K986P and V987P, wild type numbering). Recombinant proteins were produced in Expi293F cells (ThermoFisher) by transfection with purified DNA using the ExpiFectamine 293 Transfection Kit (ThermoFisher). Supernatants from transfected cells were harvested 3 days post transfection, and recombinant protein were purified using Ni-NTA agarose (ThermoFisher), then buffer exchanged into phosphate buffered saline (PBS) and concentrated using Amino Ultracel centrifugal filters (EMD Millipore).

**High-throughput assay using a recombinant VSV-SARS-CoV-2** Recombinant VSV-SARS-CoV-2 was produced as described.^2^ The neutralization was done as is described.^3^ Briefly, serial dilutions of patient sera, beginning with a 1:10 initial dilution were performed in 384-well plates and were incubated with 10^4^ PFU of VSV-SARS-CoV-2 common variant for 1 h at 37°C. Vero E6 cells were added to the human serum-virus complexes in 384-well plates at 2.5 × 10^3^ cells per well and incubated at 37°C for 16 h. Cells were fixed at room temperature in 4% formaldehyde and then rinsed with PBS. Cells were stained at room temperature with NucRed Live 647 (Invitrogen) for 30 min. Images were acquired using an InCell 6500 confocal imager (Cytiva) to visualize nuclei and infected cells then segmented using InCarta (Cytiva). Virus-infected cells were identified by comparing to the uninfected threshold in Spotfire (Tibco). IC_50_s were generated after logistic regression enforcing a plateau and baseline.

### Figure S1. Decreased Blood Plasmablast Formation in CID Participants on Prednisone after SARS-CoV-2 Vaccination.

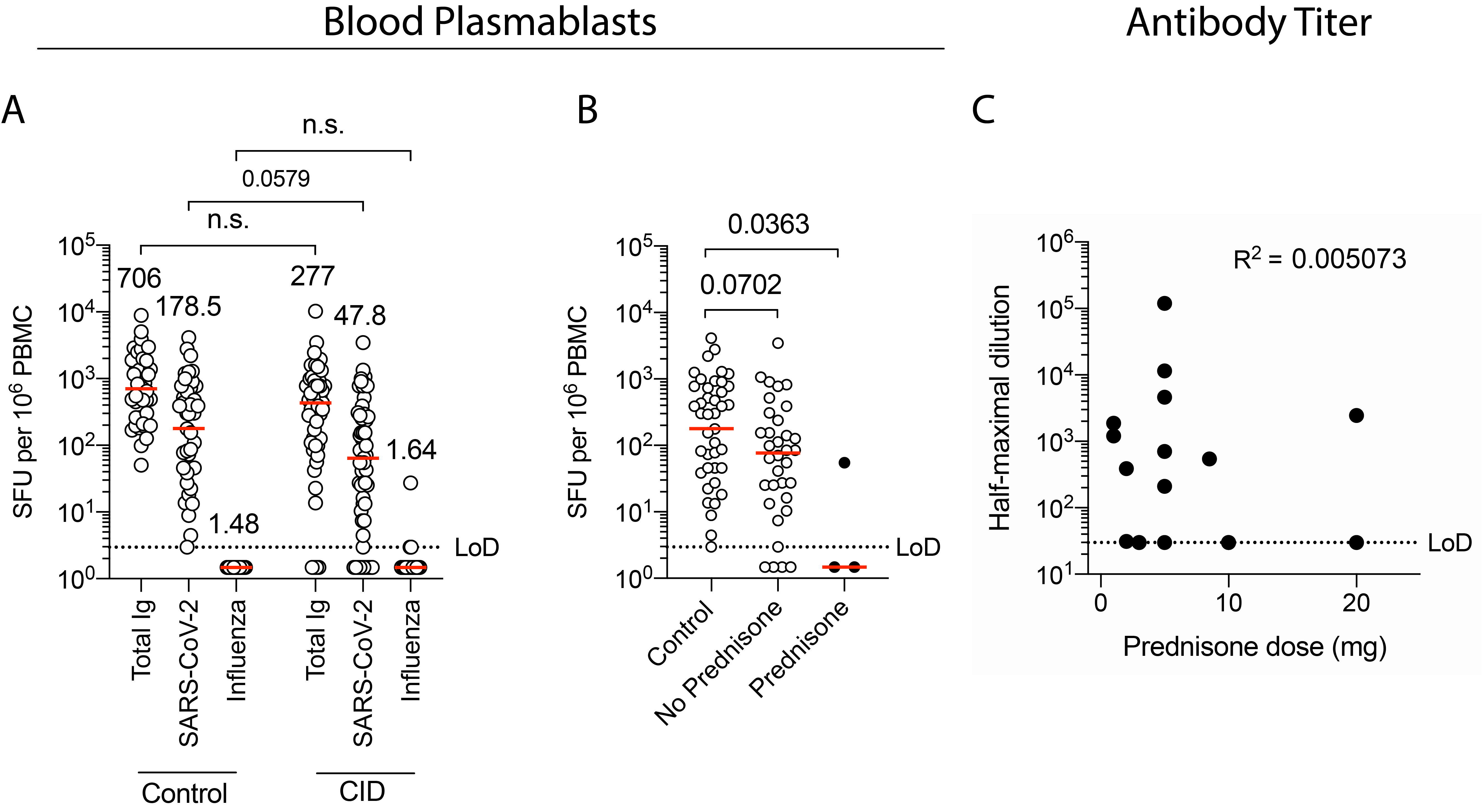

Frequency of circulating plasmablasts for immunocompetent and CID participants post-vaccination is shown in Panel A. Plasmablast frequency was measured as spot forming units (SFU) per 10^6^ PBMCs via ELISpot assay. Total IgG, SARS-CoV-2 S-binding IgG, and influenza virus vaccine-binding IgG are shown. Frequency of circulating plasmablasts for immunocompetent and CID participants on or off prednisone is shown in Panel B. A comparison of prednisone dose and anti-SARS-CoV-2 antibody titer is shown in Panel C. P values are indicated for each comparison (Kruskal–Wallis test). R-squared values (Pearson correlation) are displayed in Panel C.

### Figure S2. No Effect of Antimetabolites on Formation of Blood Plasmablasts.

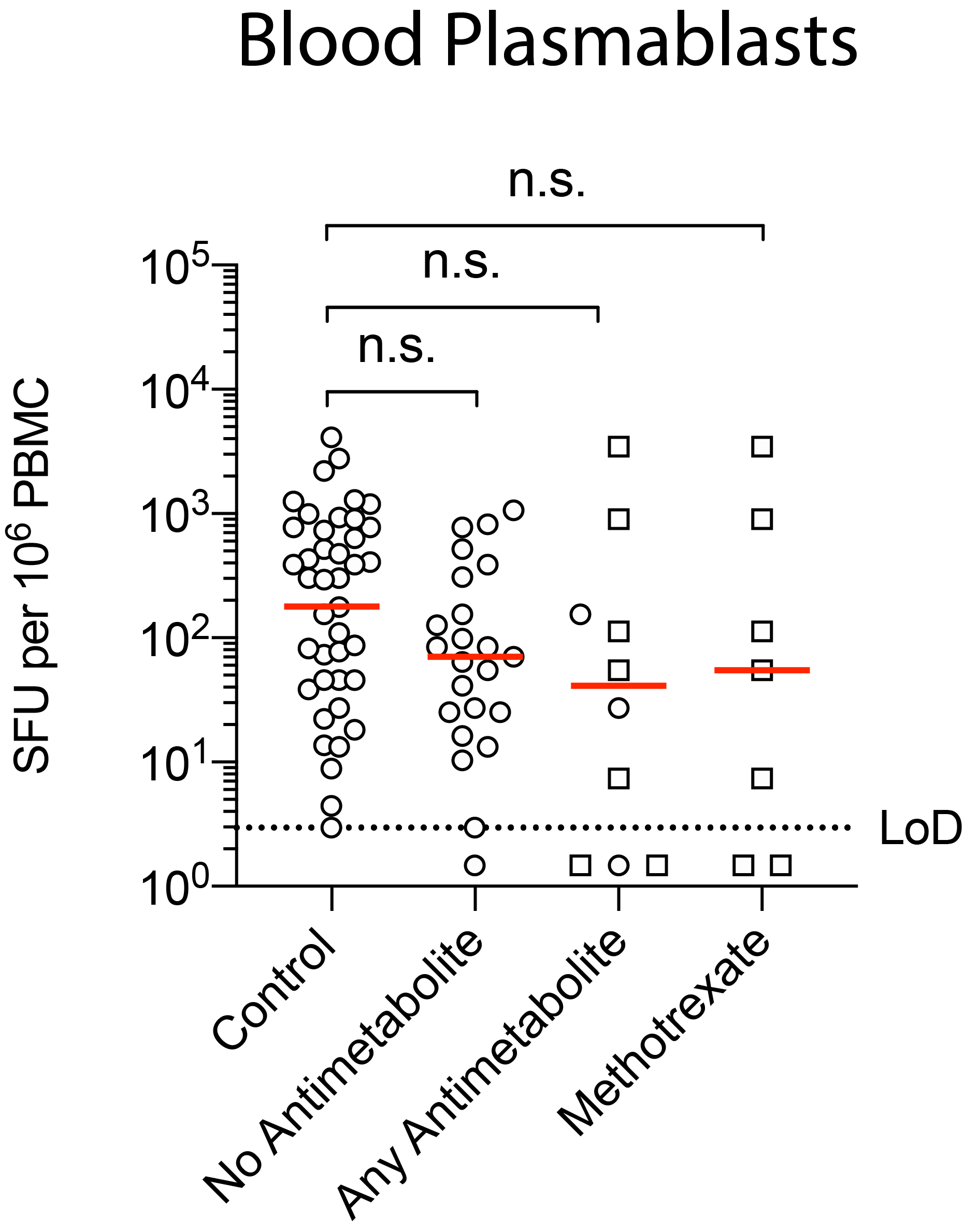

Frequency of circulating plasmablasts for immunocompetent and CID participants on or off antimetabolite or methotrexate therapy is shown. P values are indicated for each comparison (Kruskal–Wallis test).

### Figure S3. Immunogenicity of mRNA-based SARS-CoV-2 Vaccination for Each Major Therapy Class.

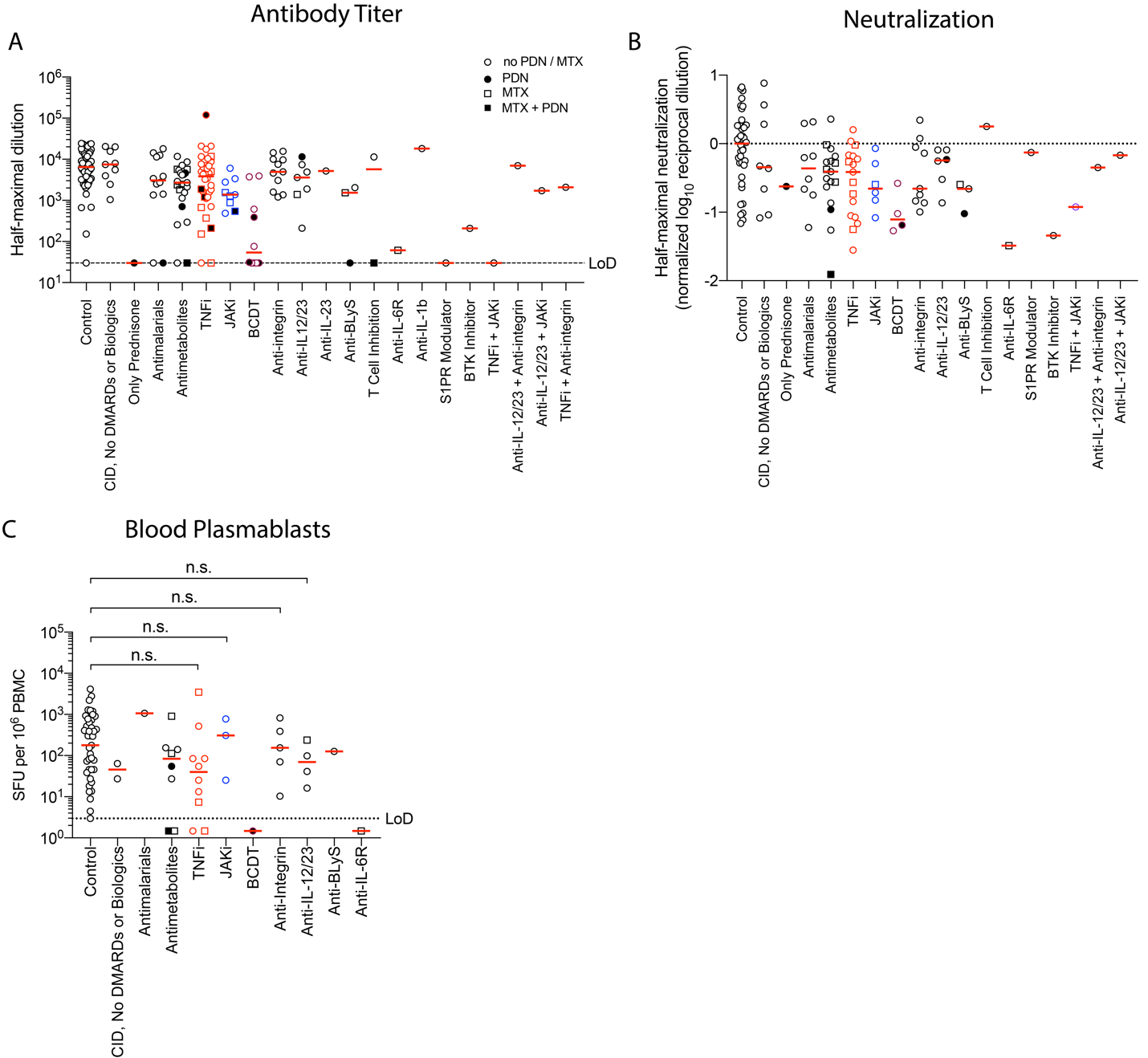

Quantification of circulating anti-spike IgG for immunocompetent (control) and CID participants separated by medical therapy: taking no DMARDs or biologic therapy, glucocorticoids alone, antimalarials, antimetabolites, TNF inhibitors (TNFi), JAK inhibitors (JAKi), B cell depletion therapy (BCDT), integrin inhibitors, IL-12/23 inhibitors, IL-23 inhibitors, BLyS inhibitors, T cell inhibitors (CTLA4-Ig), IL-6R inhibitors, IL-1b inhibitors, sphingosine-1-phosphate receptor (S1PR) modulator, Bruton’s tyrosine kinase (BTK) inhibitor, or the indicated combination therapy. Combined use with prednisone (PDN) is denoted by solid fill and with methotrexate (MTX) is denoted by square symbols. Neutralization for immunocompetent and CID participants separated by medication use in the St. Louis cohort are shown in Panel B. Frequency of circulating plasmablasts for immunocompetent and CID participants separated by medication use is shown in Panel C. P values are indicated for each comparison (Kruskal–Wallis test).

### Figure S4. Immunogenicity of mRNA-based SARS-CoV-2 Vaccination Gradually Returns after B Cell Depletion Therapy.

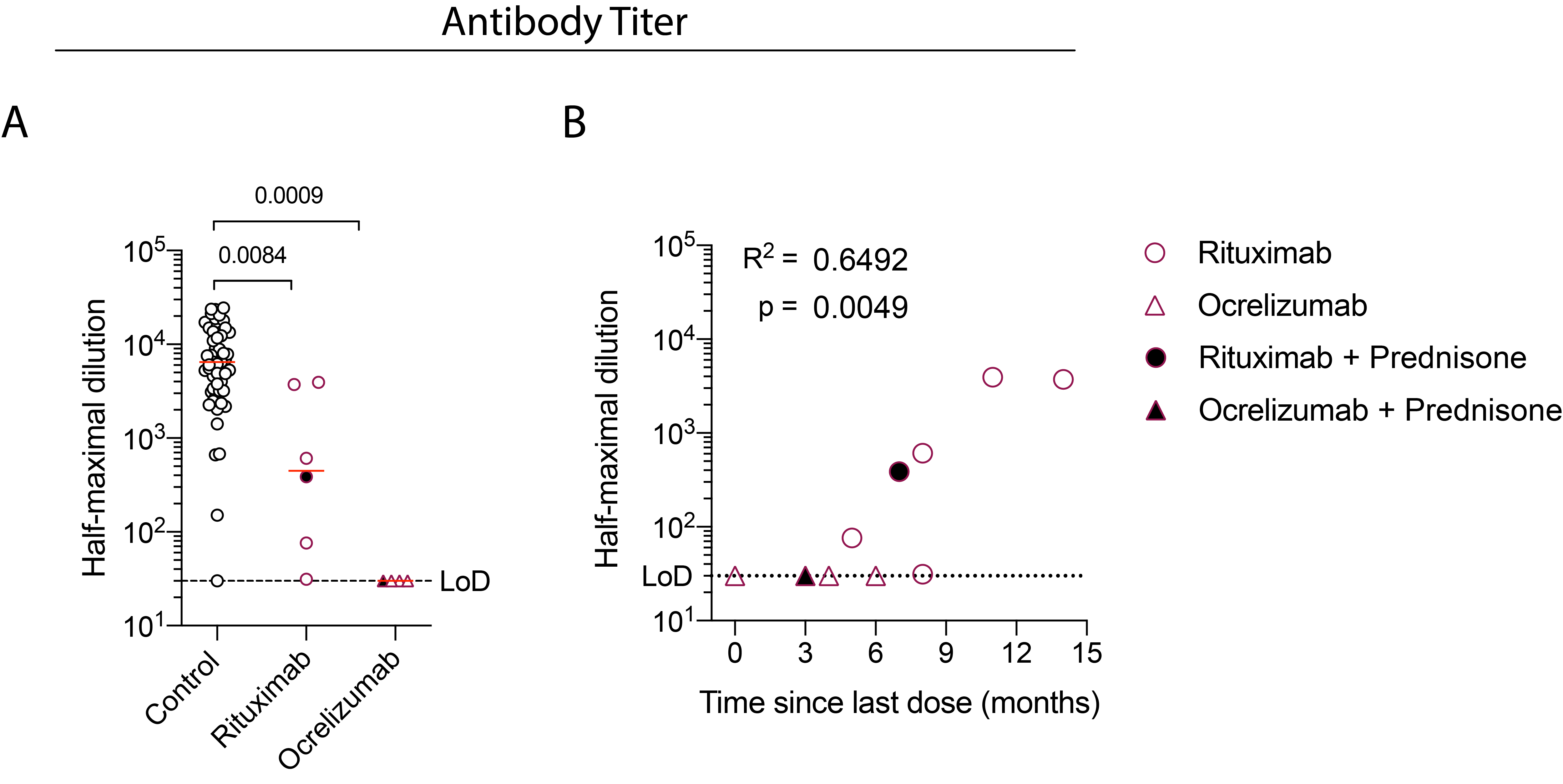

A decrease in anti-S IgG for CID participants on either rituximab or ocrelizumab is shown in Panel A. Increasing anti-S IgG post-vaccination for CID participants with increased time since last BCDT is shown in Panel B. Solid fill represents concurrent prednisone use. P values are indicated for each comparison (Kruskal–Wallis test) in Panels A and B. R-squared and P values (Pearson correlation) are displayed in Panel C.

### Figure S5. Multivariate Regression Identifies BCDT, Prednisone, JAKi, and Antimetabolites as Specific Agents that Negatively Impact Immunogenicity of SARS-CoV-2 mRNA-based Vaccination.

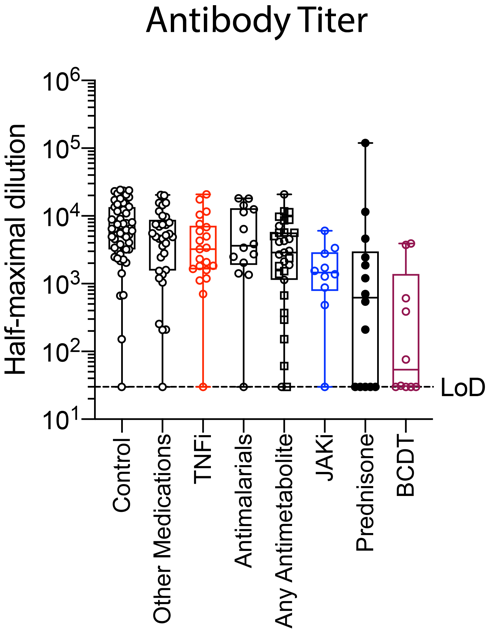

Anti-S IgG titers for immunocompetent and CID participants are shown. CID participants are grouped based on medication use and ordered based on progressive negative impact on humoral response as identified by a multivariate regression analysis.

### Table S1. Immunocompetent Control versus CID Participant Characteristics.

| **Characteristics** | | **Control (N = 53)** | **CID (N = 133)** |
| --- | --- | --- | --- |
| Age, years | |  |  |
|  | Mean ± SD | 43.4 ± 14.1 | 45.5 ± 16 |
| Age category — no. of participants (%) | |  |  |
|  | <65yr | 46 (86.8) | 114 (85.7) |
|  | ≥65yr | 7 (13.2) | 19 (14.3) |
| Gender — no. of participants (%) ^‡^ | |  |  |
|  | Female | 29 (54.7) | 99 (74.4) |
|  | Male | 24 (45.3) | 34 (25.6) |
| Hispanic or Latinx ethnicity – no of participants (%) ^‡^ | |  |  |
|  | Hispanic or Latinx | 4 (7.5) | 6 (4.5) |
|  | Not Hispanic or Latinx | 49 (92.5) | 127 (95.5) |
| Race or ethnic group — no. of participants (%) ^‡^ | |  |  |
|  | White | 42 (79.2) | 117 (88) |
|  | Asian | 7 (13.2) | 9 (6.8) |
|  | Black or African American | 1 (1.9) | 4 (3) |
|  | Other | 3 (5.7) | 3 (2.3) |

^‡^ Gender, Race or ethnic group was reported by the participant.

### Table S2. Post-vaccine Titer by CID Medication Group vs Other^1^ or No Medications in CID Participants

| **Medication group^2^** | **N** | **Fold Reduction in Titer (95% CI)** | **P-value^3^** |
| --- | --- | --- | --- |
| B-cell depleting therapy | 10 | 35.7 (9.3 – 138.0) | **4.10E-06** |
| Prednisone | 14 | 10.7 (2.5 – 45.2) | **0.0019** |
| JAK inhibitors | 10 | 4.5 (1.6 – 12.4) | **0.0047** |
| Antimetabolites | 31 | 1.8 (0.81 - 4.2) | 0.14 |
| Antimalarials | 14 | 1.5 (0.54 – 4.1) | 0.44 |
| TNF inhibitors | 21 | 1.0 (0.92 - 3.2) | 0.94 |

^1^Other Medications include NSAIDs, sulfasalazine, anti-integrin therapy, anti-IL-12/23 therapy, and ibrutinib.

^2^Each medication group excludes patients in the group(s) above.

^3^Tobit regression of log_10_(post-vaccine titer) left-censored at limit of detection log_10_(30), adjusting for log_10_(pre-vaccine titer), age, and gender.

### Table S3. Impact of the Number of Immunosuppressive Medications on the Immunogenicity with SARS-CoV-2 Vaccine Among Patients with CID

| **Within AID patients** | **Variable** | **Coefficient^2^ (log_10_ titer)** | **% change**  **titer** | **Lower 95 CI % change titer** | **Upper 95 CI % change titer** | **P-value**^1^ |
| --- | --- | --- | --- | --- | --- | --- |
| Including patients on prednisone | Number of medications | -0.236 | -41.9% | -58.1% | -19.4% | **0.0013** |
|  | Age | -0.0070 | -1.6% | -3.7% | 0.5% | 0.14 |
|  | Female gender | 0.310 | 104.2% | -8.4% | 355.0% | 0.080 |
| Excluding patients on prednisone | Number of medications | -0.0900 | -18.7% | -45.7% | 21.7% | 0.31 |
|  | Age | -0.0077 | -1.8% | -3.8% | 0.3% | 0.10 |
|  | Female gender | 0.2067 | 60.9% | -25.4% | 247.3% | 0.22 |
| **Within those on multiple medications** | |  |  |  |  |  |
| On prednisone versus not on prednisone | | -0.7729 | -83.1% | -95.2% | -41.0% | **0.0061** |

^1^Tobit regression of log_10_(post-vaccine titer) left-censored at limit of detection log_10_(30), adjusting for log_10_(pre-vaccine titer), number of immunosuppressive medications, age, and gender, comparing those on prednisone vs not.
